# Deep continual multitask out-of-hospital incident severity assessment from changing clinical features

**DOI:** 10.1101/2024.02.20.24303094

**Authors:** Pablo Ferri, Carlos Sáez, Antonio Félix-De Castro, Purificación Sánchez-Cuesta, Juan M García-Gómez

## Abstract

When developing Machine Learning models to support emergency medical triage, it is important to consider how changes over time in the data can negatively affect the models’ performance. The objective of this study was to assess the effectiveness of novel Deep Continual Learning pipelines in maximizing model performance when input features are subject to change over time, including the emergence of new features and the disappearance of existing ones. The model is designed to identify life-threatening situations, predict its admissible response delay, and determine its institutional jurisdiction. We analyzed a total of 1 414 575 events spanning from 2009 to 2019. Our findings demonstrate important performance improvements, up to 4.9% in life-threatening, 18.5% in response delay and 1.7% in jurisdiction, in absolute F1-score, compared to the current triage protocol, and improvements up to 4.4% in life-threatening and 11% in response delay, in absolute F1-score, respect to non-continual approaches.

## 1 Introduction

Out-of-hospital emergency medical triage presents a complex challenge, involving fast-paced decisions with considerable uncertainty, where errors can have fatal consequences. To aid dispatchers and reduce variability among professionals, emergency medical dispatch centers provide clinical guidelines, collectively known as clinical protocols [1]. These protocols include well-known systems such as the Manchester Triage System [2], the Canadian Triage Scale [3] or the Emergency Severity Index [4], which share a common structural arrangement as decision trees with clinical queries and branching pathways leading to a terminal node specifying the assigned priority level for the incident.

Within the domain of out-of-hospital emergency medical triage in the Valencian Community (Spain), an in-house triage protocol was conceived by experts within the Health Services Department (HSD) of this region. Initially inspired by the Manchester Triage System, the protocol underwent iterative adaptations over time, drawing upon the insights and expertise of coordinator physicians. Consequently, the protocol is hierarchical, featuring queries linked to distinct branches. The responses to these queries correspond to values attributed to structured clinical variables, culminating in final leaf nodes that are related to specific priority levels.

Nonetheless, the phenomenon of distributional drifts [5, 6] manifests over time. In the context of healthcare processes and medicine, these distributional variations are intrinsic [7–10], and out-of-hospital emergency medical triage processes in the Valencian region are no exception [11]. Focusing on this specific context, the occurrence of these shifts is attributed to a multitude of factors. Foremost, it is imperative to underscore the alteration in the information system during 2013, which engendered substantial shifts in protocols, personnel, and emergency coordination. Furthermore, changes in telephone operators, targeted training initiatives and updates to clinical variables via the evolution of the in-house triage protocol, have collectively exerted their impact over time.

In the context of the Valencian region, a deep multitask ensemble model named DeepEMC^2^ was developed to support incident severity assessment processes within this area [12]. This model is constituted by four main deep neural networks, each one specialized on a specific data type: the *Context network*, focused on demographics and circumstantial data; the *Clinical network*, designed to deal with the clinical features derived from the protocol; the *Text network*, which considers the free text data written by the dispatcher; and the *Ensemble network*, which combines the inner representations of the previous networks to offer severity assessment. DeepEMC^2^, the global network resulting from the combination of the later four, showed promising results when compared with the in-house triage protocol of the Valencian region, providing performance enhancements of 12.5%, 17.5%, and 5.1% when assessing the life-threatening level, the admissible response delay and the jurisdiction of incoming incidents, in terms of absolute macro F1-score. Hence, its implantation could derive in a huge positive impact over patient well-being and health services sustainability.

However, due to data availability reasons, the data considered to train and evaluate these multiple networks covered the period from 2009 to 2012. As previously discussed, when developing a Machine Learning-based system for providing triage decision support, it is crucial to consider the dataset shift phenomenon that naturally occurs over time [11, 13, 14]. In the context of the Valencian region, shifts associated with the appearance and disappearance of clinical features over time pose a tough challenge that, if unaddressed, could harm model performance through time.

Given the potential negative performance impacts resulting from the changing environment, it is imperative to incorporate mechanisms to mitigate the adverse effects arising from these shifts. Hence, within this study, we have designed novel deep architectures for the *Clinical network* along with novel Deep Continual Learning pipelines [15, 16], aiming to face the changing clinical features challenge over time, while maximizing model performance. After their design, we have implemented and applied them to multiple temporal sets of out-of-hospital emergency medical data from the Valencian region, to assess their effectiveness. Despite the existence of previous works addressing the training and deployment of Machine Learning models in the context of medical data with temporal dataset shifts [11, 13, 14, 17–19], to our knowledge, this is the first study facing evolving clinical features within the domain of out-of-hospital emergencies.

## 2 Materials

We considered a total of 1 414 575 independent out-of-hospital emergency medical incidents from the HSD of the Valencian region, compiled from 2009 to 2019, excluding 2013—since the emergency information system changed during that year. Data usage was approved by the Institutional Review Board of the HSD. No information that may disclose the identity of the patient was kept for any of the analyses.

The data employed in these studies encompassed both during-call and after-call data. During-call data were recorded during the emergency medical call and included clinical tree variables and values associated with the in-house decision tree. Some examples of possible clinical features sets associated each one to a different incident are: 1) “Previous trauma: no; Shortness of breath: yes; Nasal congestion: no” and 2) “Active arrhythmia: yes; History: cardiac pathology; Dizziness: yes; Incident location: public road/street”. These data were used at inference time as input for the prediction. On the other hand, after-call data were recorded at a time after the call. They include physician diagnosis, hospitalizations, urgency stays, maneuvers, and procedures the patient underwent. After-call data were used offline—i.e., not in prediction time—to infer the output variables of the predictive model: if the emergency event implied or not a life-threatening situation, which was the admissible response delay—undelayable, minutes, hours, days—and if the event was jurisdiction of the emergency system or primary care.

As stated in the previous section, the clinical tree features have undergone significant changes over time. Some features have experienced shifts in their occurrence rates, others have been entirely phased out after several years, and new features have emerged. This phenomenon is illustrated in Figure 1, which depicts the empirical frequency—specifically, the empirical square root frequency for ease of visualization—of the most common clinical features in our dataset across different time periods.

**Fig. 1.**
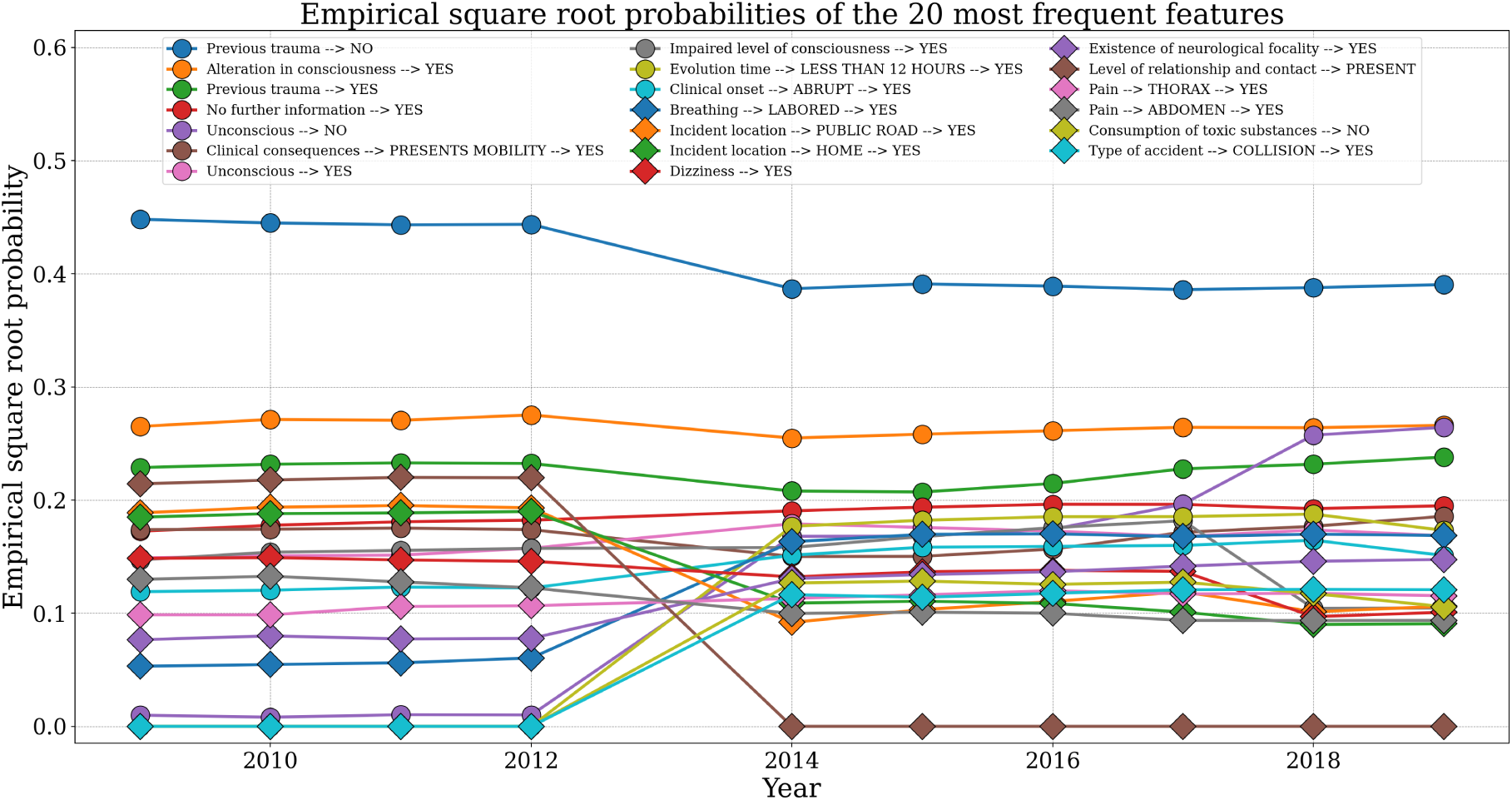
Square root of the empirical probabilities associated with the 20 most frequently occurring clinical features in our dataset.

## 3 Methods

### 3.1 Data preparation

We segmented our data to capture temporal variations while assessing overfitting. We initially partitioned our dataset into distinct time windows. The first window comprised data solely from the years spanning from 2009 to 2012 and, subsequently, each ensuing time window corresponded to a specific year (2014-2019). This partitioning strategy was selected due to forthcoming architectural and training variations within the original *Clinical network*. This network was trained collectively on the entire 2009-2012 data batch, rather than in a year-by-year manner. Thus, if we intend to evaluate the impact of architectural modifications, it is imperative to ensure the use of consistent data sets for performance metric comparisons.

Following this initial partition, another division was performed. For each time window, a training and test split was conducted, allocating 80% for training and 20% for testing. Subsequently, the training set was divided into a pure training subset and a validation subset, with proportions 70% and 30%, respectively. The validation subset was exclusively employed for hyperparameter tuning, with no inclusion of cases from the test set.

Subsequently, we transformed the categorical clinical features into indexes. This conversion was necessary to enable the subsequent utilization of an Embedding Layer [20], which will map every index to a dense vector in our models. Additionally, we undertook padding and truncation operations to ensure a consistent sequence length, thereby fastening training processes.

Finally, with respect to the labels, they were encoded through one-hot encoding. As such, from the life-threatening label, two variables were derived. Likewise, four variables originated from the admissible response delay label and an additional two from the emergency system jurisdiction label.

### 3.2 Deep neural network design

Considering the outcomes detailed in [12], where Deep Learning models exhibited superior performance compared to other Machine Learning approaches such as Random forest [21] or Gradient boosting [22], the focus in this work remains on models of a similar nature. However, the present study excludes the adoption of recurrent architectures as the original *Clinical Network*, since we aim to build an order-invariant model, able to generate consistent predictions even when clinical features are presented in varying orders and hence, being robust to this type of shifts. In addition, this model has to be able to handle the challenge posed by the emergence and disappearance of novel features over time.

In the next section, we introduce the model that we developed to align with these specific requirements. Due to its inherent characteristics, we have named this model the *Clinical Invariant Network*, denoted as CliInvNet for brevity.

#### 3.2.1 Clinical Invariant Network

The *Clinical Invariant Network* comprises a multitask [23] deep neural network, constituted by two main components: the Indexes Encoder and the Multitask Classifier. The Indexes Encoder, serving as the network’s hard parameter sharing element, forms its core. Meanwhile, the Multitask Classifier contains distinct branches, each associated with a specific label. These branches are responsible for computing predicted scores for the various classes within each label.

Focusing on the Indexes Encoder, we constructed it with an initial Embedding Layer [20]. This layer facilitates the mapping of clinical variables, expressed as indexes, into dense vector representations, a significantly more efficient alternative to one-hot encodings. Moreover, this Embedding Layer enables the accommodation of novel features over time. We achieve this by pre-allocating a substantial number of entries within the corresponding lookup matrix without impacting subsequent architectural elements. Following the Embedding Layer, an Adaptive Average Pooling block [24] was employed. This component serves to aggregate the representations of all features within an observation into a singular representation. This functionality allows the network to accommodate varying numbers of features per entry. Additionally, the Adaptive Average Pooling Layer endows the network with order-invariant capabilities, preserving results even with altered feature orders. Following this, multiple dense blocks were introduced, each encompassing a Fully Connected Layer [25], Layer Normalization [26], a GELU activation function [27], and a Dropout Layer [28] to counteract neuron co-adaptation.

The Multitask Classifier, responsible for incorporating task-specific components into the architecture, consists of three branches. Each branch contains several dense blocks, culminating in an output block. These output blocks consist of a Fully Connected Layer followed by a Softmax activation function. Illustrated in Figure 2, the main architecture of CliInvNet is visually represented.

**Fig. 2.**
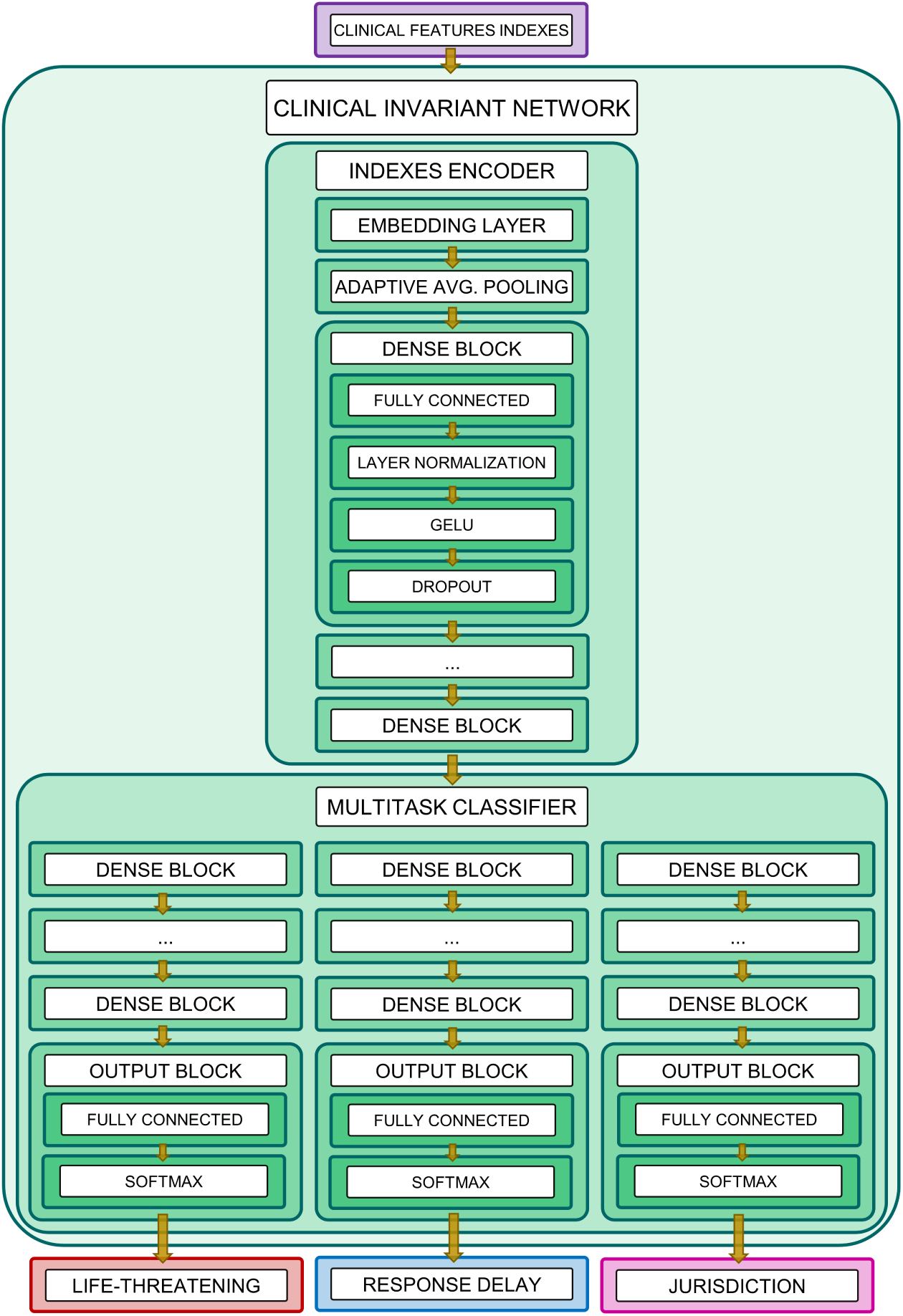
Clinical Invariant Network architecture. It is a deep multitask neural network composed of two primary components: the Indexes Encoder and the Multitask Classifier. The Indexes Encoder transforms input clinical variables, encoded as indexes, into deep continuous embeddings. The Multitask Classifier then utilizes these embeddings to make predictions. It assesses the probability of a life-threatening event, categorizes the admissible response delay at various levels, and determines whether the incident falls under the jurisdiction of the emergency system.

### 3.3 Parameter tuning

Concerning the parameter tuning process, we used the AdamW [29] optimizer. The feeding paradigm followed was a mini-batch training approach [30], while the loss function considered was the soft F1-score [31]. To further enhance our training, we incorporated a cosine annealing learning rate scheduler, which aligns aptly with deep transfer learning scenarios [32].

Likewise, layers featuring ReLU activation functions were initialized using Kaiming initialization [33], whereas layers incorporating the softmax activation function were initialized with Xavier’s initialization [34].

### 3.4 Continual Learning

In this section, we present Continual Learning strategies specifically designed to support the CliInvNet’s adaptation across different time windows. It is important to emphasize a crucial distinction at this point: the difference between approaches that focus on defining how the evolving clinical features should be represented and updated over time windows—strategies for handling changing feature domains—and procedures concerned about how CliInvNet’s parameters should be updated over the different time windows—strategies to update parameters over time windows.

#### 3.4.1 Strategies for handling changing feature domains

##### Static domain

The static domain strategy involves using the feature identifier-to-index conversion map from the original Clinical Network (CliNet) within DeepEMC^2^. This map is fixed after the initial time window and does not receive updates thereafter. If new variables arise that correspond to the 2009-2012 data batch, they are mapped to a known clinical feature. Conversely, features appearing over time without any prior correspondence are assigned to the index representing unknown or infrequent nodes within the CliNet. As a result, the number of active entries in the CliInvNet’s Embedding Layer stays constant across all time windows. However, the values of these dense representations vary over time as the model incorporates new data.

##### Dynamic domain

The dynamic domain strategy is based on the recurrent update of the feature identifier-to-index map with each new time window. We establish a frequency threshold, mirroring the one employed in the CliNet, to discern when a feature qualifies as infrequent. Such features are then either assigned to the unknown index or mapped to a distinct integer designated solely for that feature. Across the series of time windows, we monitor and revise the cumulative absolute frequency of each feature’s occurrences. This iterative process facilitates the emancipation of features that were initially mapped to the unknown integer, permitting their adaptation in subsequent time windows and preventing them from becoming stagnant. Hence, the number of active entries of the Embedding Layer of the CliInvNet varies over the time windows as long as the cumulative clinical variable frequency surpasses the required threshold. In addition, the value of those enabled dense representations vary over time as the model learns from new data.

##### Predefined domain

The predefined domain strategy employs a predefined embedding matrix derived from a large pre-trained natural language processing model. Specifically, for this work, we selected the ALBERT model [35] pretrained on a Spanish corpus [36]. This choice is motivated by the fact that our dataset contains clinical variables originally in Spanish. Additionally, the dimensionality of the ALBERT model’s embeddings closely matches that of the embeddings used in the static and dynamic approaches, enabling effective comparisons across these strategies.

Under this feature domain approach, the structured clinical variables are transformed into an unstructured natural language processing representation. Subsequently, we apply subword tokenization to the unstructured clinical data, breaking down the text into smaller, meaningful subtokens. After subword tokenization, we utilize the embedding matrix derived from the pretrained ALBERT model. This matrix allows us to obtain stable numerical representations for each subtoken. By leveraging a pretrained NLP model like ALBERT, we harness its capacity to capture semantic and contextual information from the clinical features in text format, thus enhancing the quality of our embeddings.

The predefined approach is intended to maintain stability through the consistent use of the ALBERT embedding matrix across all time windows. This ensures that the numerical representations for clinical variables remain robust and consistent, even as the model learns from new data.

We illustrate in Figure 3 a schematic representation of the three strategies for handling evolving feature domains proposed in our work.

**Fig. 3.**
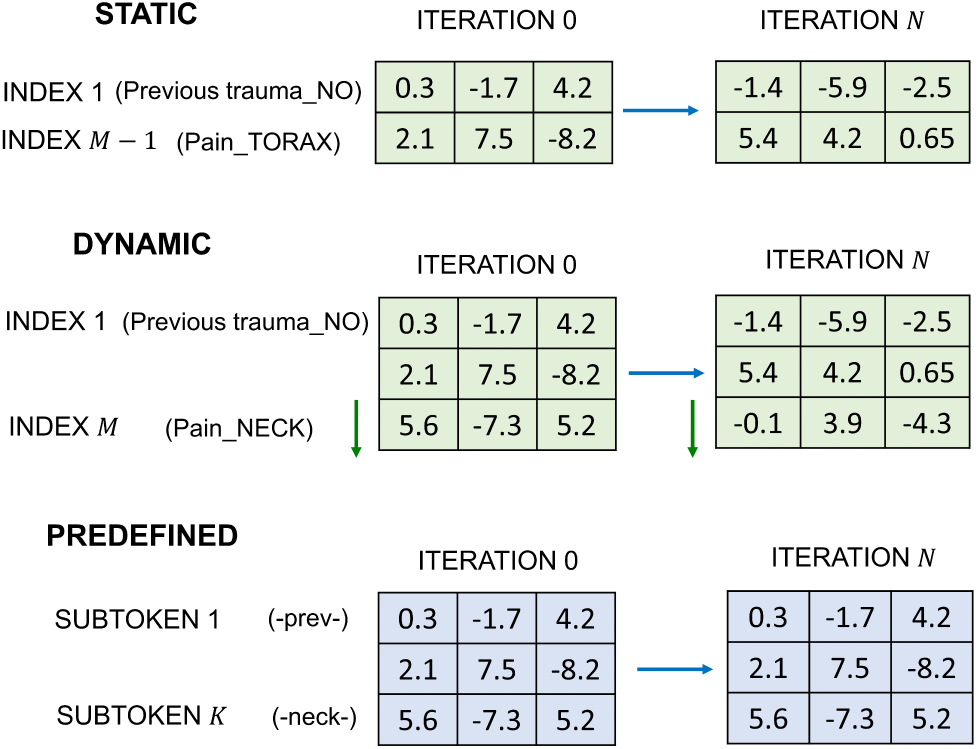
Depiction of the three main strategies for handling changing feature domains proposed in our work: the static domain strategy, the dynamic domain strategy, and the predefined domain strategy. The numbers are dense continuous vectors representing entries associated with the Embedding Layer of the *Clinical Invariant Network*.

#### 3.4.2 Strategies to update parameters over time windows

##### From scratch

The from-scratch approach involves exclusively utilizing data from the current time window, necessitating the initialization of a new model and training it anew on each occasion. This approach may appear to be less advantageous, given that it incorporates a substantially smaller dataset compared to the majority of Continual Learning strategies. However, in scenarios where there are pronounced dataset shifts over time, this approach may indeed be prudent. By eschewing the integration of noise from previous time windows into the current one, it emerges as a sensible option.

##### Continual fine-tuning

The continual fine-tuning strategy entails retraining the model exclusively with data from the most recent time window. For each new time window, the model begins with the adjusted weights from the previous training session’s conclusion. This approach is adopted because it strikes an effective balance: it retains some past information since the model weights are not randomly initialized, yet it primarily facilitates the transfer of forward knowledge, thereby avoiding an over-reliance on past time windows.

##### Cumulative

In this strategy, data from the current time window is combined with data from all preceding time windows. Consequently, the volume of data employed for training expands with each new time window. This augmentation in data utilization brings about heightened computational demands and memory requirements. However, it offers the advantage of retaining a comprehensive record of previous data patterns. As a result, the model stands to benefit from a data accumulation standpoint—an advantageous attribute, given that Deep Learning models tend to exhibit enhanced performance with a larger pool of available data.

### 3.5 Evaluation

To evaluate the performance of each deep continual pipeline and determine the one best suited for severity assessment support over time, we calculated the F1-score associated with each severity label for every pipeline. Specifically, we computed the F1-score for the positive class of the “life-threatening” label (i.e., the “life-threat” class) and the “jurisdiction” label (i.e., the “emergency system jurisdiction” class). For the “admissible response delay” label, we calculated the F1-score using macro-averaging, as we cannot designate a reference class among the four classes.

To assess the real out-of-sample effect inherent to dataset shifts, we computed these metrics for each time window, but considering the training of the model up to the previous time window. This way, we can understand how model performance diminishes when applied to novel incoming data, which may exhibit variations.

Then, we averaged and studied the performance for each feature domain and parameter updating strategy over the different time windows to gain a better understanding of the effect of each approach. Additionally, we obtained non-parametric 95% confidence intervals using bootstrap resampling [37], with a total of 1000 resamples per pipeline.

Finally, we calculated the performance metrics for each time window for the in-house triage protocol of the Valencian region, as well as those for the original *Clinical network* from DeepEMC^2^. These metrics served as baselines to assess the value added by the implemented deep continual approaches.

## 4 Results

### 4.1 Life-threatening

Upon observing Figure 4 it becomes evident that all the deep continual pipelines designed in this study provide substantial value with respect to the triage protocol as well as to the original *Clinical network*, which does not consider a continual approach. Notably, most deep continual pipelines exhibit their weakest out-of-sample performance in the 2014 time window, with improvements in subsequent years. Likewise, a downward trend in performance begins in 2015, becoming more pronounced for the triage protocol and the CliNet.

**Fig. 4.**
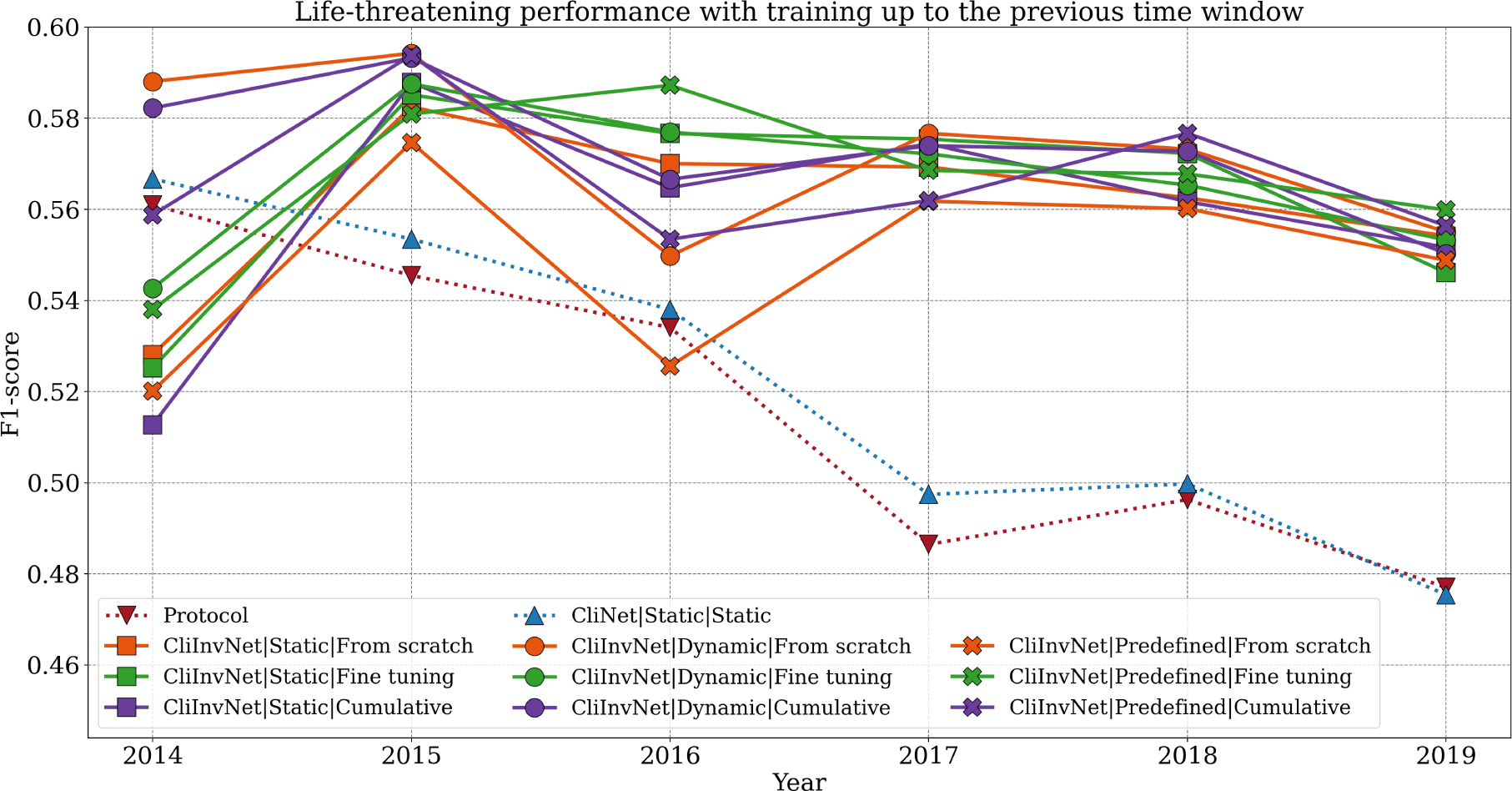
Life-threatening performance over time with training up to the previous time window for each pipeline. Abbreviations: CliNet, *Clinical network*; CliInvNet, *Clinical Invariant Network*.

From the analysis of Table 1 it is noteworthy to highlight the differences among the static, dynamic, and predefined feature domain approaches, with the dynamic approach outperforming the others. Regarding parameter updating strategies, the outcomes for the fine-tuning and cumulative approaches are closely matched, whereas the from scratch approach yields inferior results.

**Table 1.** Average F1-score values for life-threatening performance with training up to the previous time window for each deep continual pipeline tested. Non-parametric 95% confidence intervals for each average value are provided between brackets.

| Parameter updating | Feature domain |  |  |  |
| --- | --- | --- | --- | --- |
|  | Static | Dynamic | Predefined | Mean |
| From scratch | 0.561 [0.558, 0.565] | 0.573 [0.569, 0.576] | 0.549 [0.545, 0.552] | 0.561 [0.557, 0.564] |
| Fine-tuning | 0.563 [0.56, 0.567] | 0.566 [0.563, 0.57] | 0.567 [0.564, 0.57] | **0.566 [0.562, 0.569]** |
| Cumulative | 0.559 [0.555, 0.562] | 0.573 [0.57, 0.576] | 0.567 [0.563, 0.57] | **0.566 [0.563, 0.57]** |
| Mean | 0.561 [0.558, 0.565] | **0.571 [0.567, 0.574]** | 0.561 [0.557, 0.564] | 0.564 [0.561, 0.568] |

### 4.2 Admissible response delay

From an examination of Figure 5, it is clear that all the deep continual pipelines designed in this study provide substantial value with respect to the triage protocol as well as to the original *Clinical network*, which does not consider a continual approach. The weakest out-of-sample performance of the deep continual pipelines are achieved in 2014, from which performance highly increases. This contrasts with the in-house protocol and the CliNet, which both exhibited better performance in 2014, followed by a gradual decline thereafter.

**Fig. 5.**
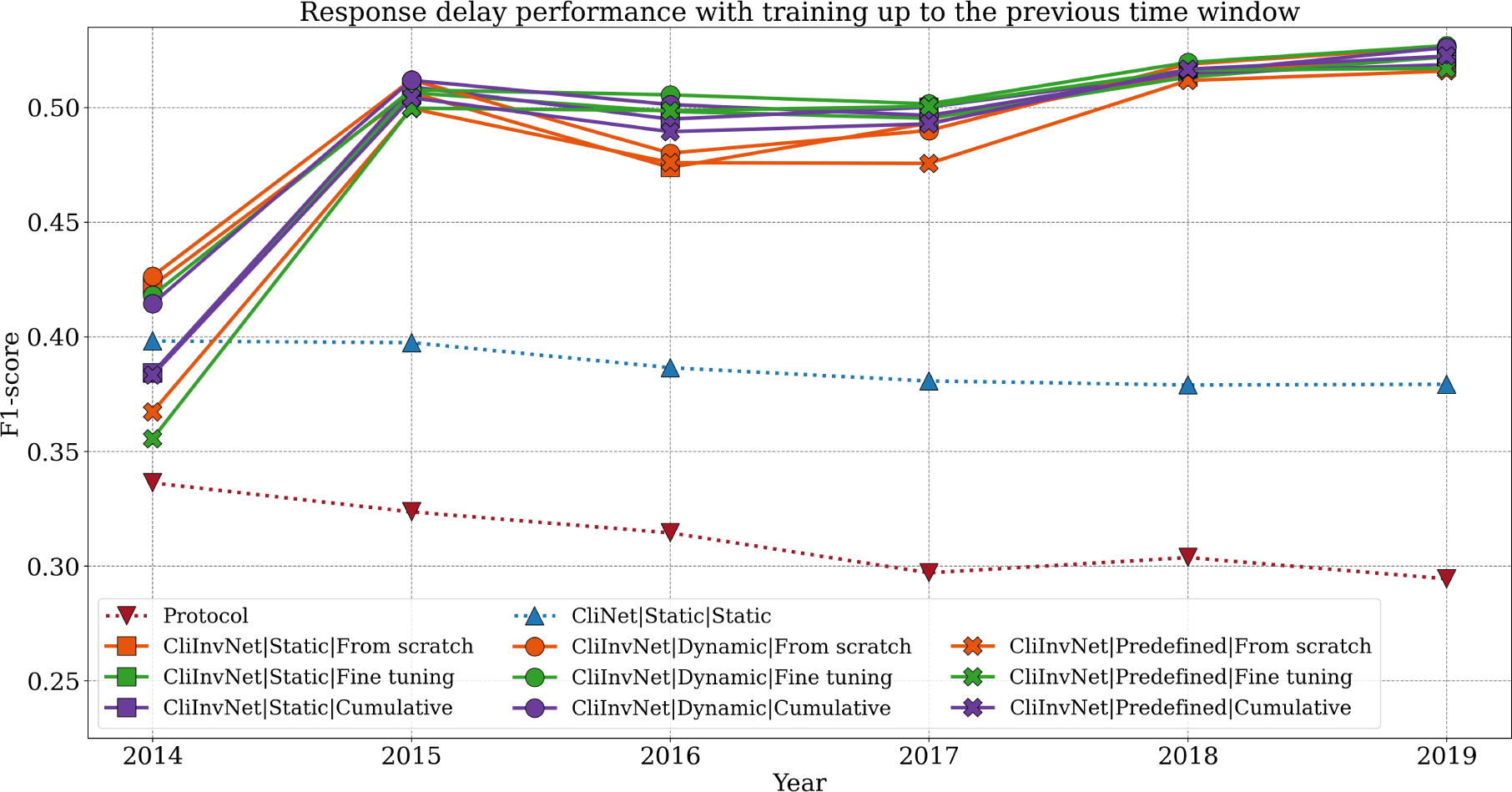
Admissible response delay performance over time with training up to the previous time window for each pipeline. Abbreviations: CliNet, *Clinical network*; CliInvNet, *Clinical Invariant Network*.

After observing Table 2 it is evident that the dynamic feature domain approach presents a more favorable behavior than the static and predefined feature domain paradigms. Regarding parameter updating strategies, the comparison reveals that the fine-tuning strategy and the cumulative approach yield the most favorable results, followed by the from-scratch strategy.

**Table 2.**
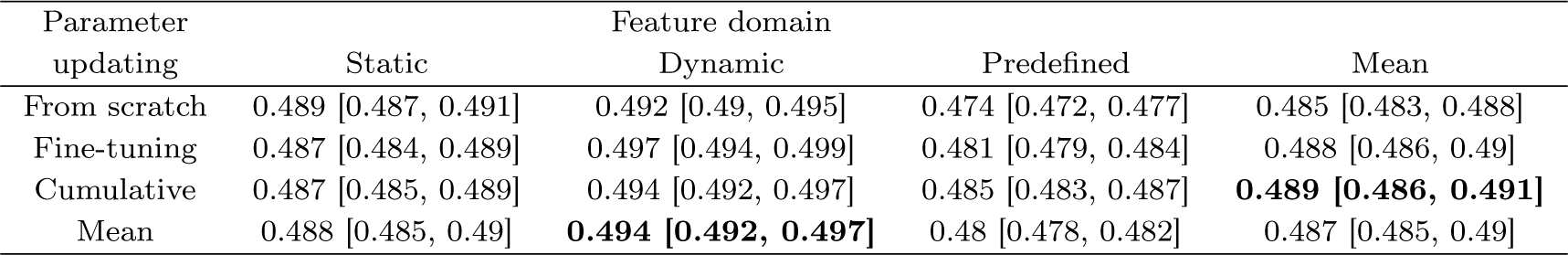
Average macro F1-score values for admissible response delay performance with training up to the previous time window for each deep continual pipeline tested. Non-parametric 95% confidence intervals for each average value are provided between brackets.

### 4.3 Emergency system jurisdiction

Upon observing Figure 6, it is inferred that the deep continual pipelines provide a notable improvement in performance over time compared to the in-house triage protocol. While some pipelines slightly outperform the outcomes of CliNet, the performance of CliNet generally aligns with the trend set by the deep continual pipelines, without significant deviations. Although some pipelines experience noticeable performance declines in 2014, the overall behavior remains quite stable with only minor fluctuations.

**Fig. 6.**
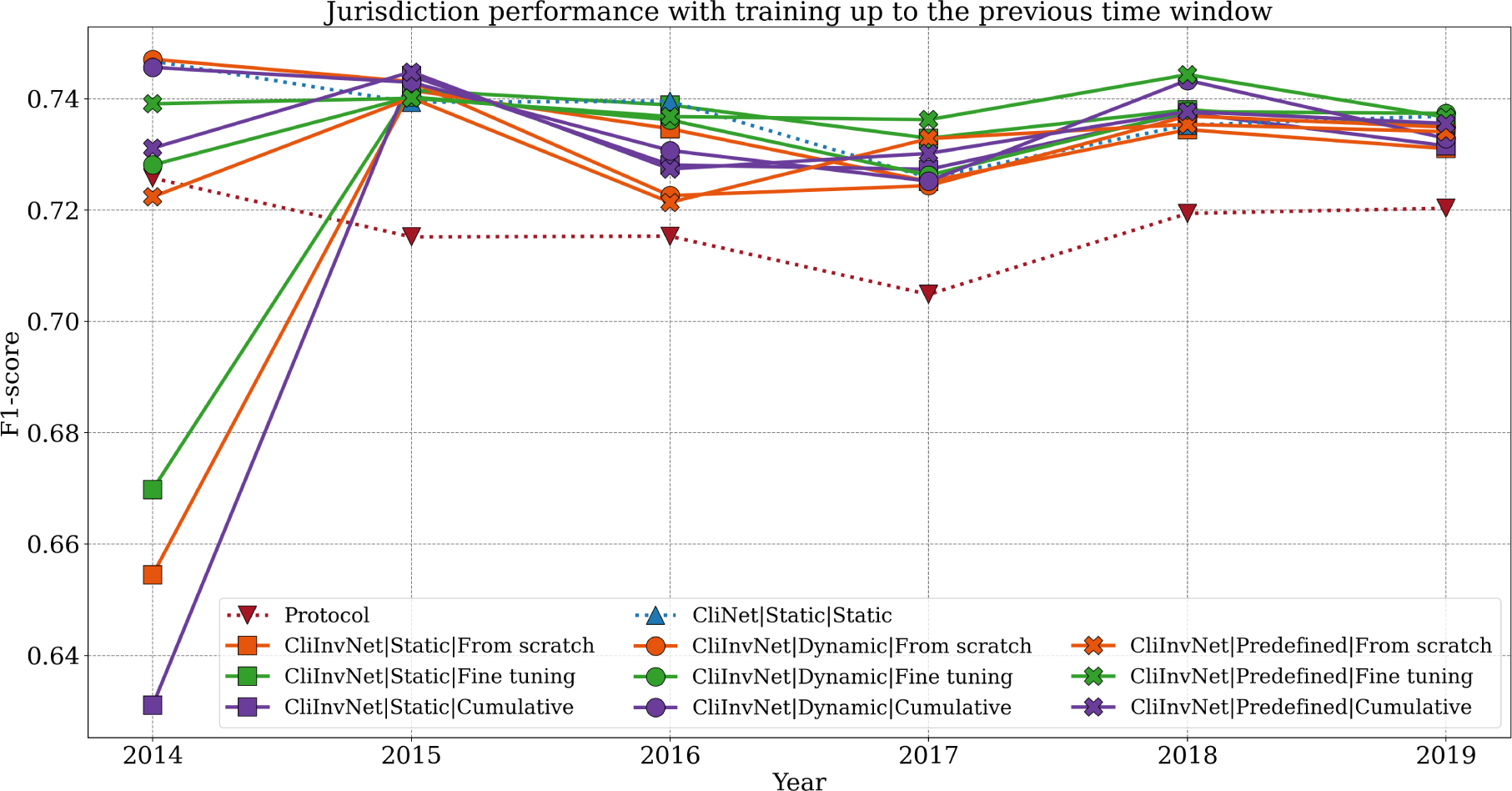
Emergency system jurisdiction performance over time with training up to the previous time window for each pipeline. Abbreviations: CliNet, *Clinical network*; CliInvNet, *Clinical Invariant Network*.

From the analysis of Table 3 it is observed that both the dynamic and predefined feature domain approaches surpass the performance of the static approach, with the dynamic and predefined methods exhibiting similar levels of performance. When assessing different strategies for updating model parameters over time, fine-tuning stands out as the most effective strategy.

**Table 3.** Average F1-score values for emergency system jurisdiction performance with training up to the previous time window for each deep continual pipeline tested. Non-parametric 95% confidence intervals for each average value are provided between brackets.

| Parameter updating | Feature domain |  |  |  | Mean |
| --- | --- | --- | --- | --- | --- |
|  | Static | Dynamic | Predefined |  |  |
| From scratch | 0.720 [0.718, 0.722] | 0.735 [0.733, 0.737] | 0.731 [0.729, 0.733] |  | 0.729 [0.726, 0.731] |
| Fine-tuning | 0.726 [0.724, 0.728] | 0.734 [0.732, 0.736] | 0.739 [0.737, 0.741] |  | **0.733 [0.731, 0.735]** |
| Cumulative | 0.717 [0.714, 0.719] | 0.737 [0.735, 0.739] | 0.734 [0.732, 0.737] |  | 0.729 [0.727, 0.731] |
| Mean | 0.721 [0.719, 0.723] | **0.735 [0.733, 0.737]** | **0.735 [0.733, 0.737]** |  | 0.730 [0.728, 0.732] |

## 5 Discussion

The findings of our study underscore the advantages of employing a Deep Continual Learning approach for assessing out-of-hospital incident severity, in scenarios where clinical features evolve over time and dataset shifts occur. The deep continual pipelines significantly surpass the performance of the in-house triage protocol of the Valencian region across all three severity labels examined in this research. Moreover, these pipelines demonstrate considerable value over non-continual approaches for the life-threatening label and the admissible response delay label.

In evaluating the feature domain strategies within the context of our out-of-hospital medical emergency data, the dynamic feature domain approach emerges as particularly beneficial for predictions in the upcoming time window. Consequently, we recommend selecting this approach over the static and predefined methods.

Concerning the strategies for updating model parameters over time, continual fine-tuning is identified as the optimal choice. Although it may not always yield the best results, this strategy is prominent for its balance of effectiveness and computational efficiency. It enables substantial knowledge transfer at a manageable computational cost, unlike the cumulative approach. Moreover, by retaining partial information from past time windows during its initialization phase, it offers an advantage over the from scratch approach, making it the preferred strategy for parameter updating in our context.

The observation that the cumulative approach does not always result in optimal performance, despite utilizing a larger training data pool, can likely be linked to the challenges posed by dataset shifts. In such scenarios, adding data from earlier time windows may introduce noise, detracting from prediction performance rather than improving it. Hence, it is plausible to suggest that retraining with historical data might hinder effective knowledge transfer and that disregarding information from previous time windows could prove more beneficial.

Another point for consideration is that the *Clinical network* keeps its performance at consistent levels from 2014 to 2019 for the emergency system jurisdiction label. This could be explained by the relative ease of predicting this particular label, suggesting that despite data variations over time, these changes have not significantly impacted performance for this label since 2014.

While there are studies addressing the training and deployment of Machine Learning models in the context of medical data with temporal shifts [11, 13, 14, 17–19], it is challenging to find similar studies for out-of-hospital emergencies. Our approach is centered on changing clinical features instead of free text features as in [11]. Furthermore, although the predefined feature domain strategy shares some similarities with the domain invariant feature approach proposed by [13] and the foundational model strategy described in [18], our approach in this work differs from previous solutions. We do not rely on raw feature aggregation [17], pre-shift patient weighting [14], or parsimonious models [19]. We instead integrate various strategies to address both the variability in the feature domain and the dynamic nature of parameter updating. This approach effectively evaluates their impact and offers an appropriate solution to the issue of changing feature domains.

Finally, it must be noted that this study is part of the ongoing developments aimed at effectively implementing a supportive Machine Learning-based tool in the emergency medical dispatch center of the Valencian region. Therefore, the findings from this research will directly influence the performance of severity assessment support in this area, impacting patient well-being and the sustainability of health services.

## 6 Conclusions

Throughout time, data in healthcare and medicine naturally undergoes changes due to factors such as population evolution, the introduction of new health policies, and updates in information systems, leading to dataset shifts. These distributional changes, if unaddressed, can severely harm the performance of any Machine Learning model over time. In this work, our focus has been on the design, implementation, and evaluation of novel Deep Continual Learning pipelines centered on providing estimates of the life-threatening level, the admissible response delay, and emergency system jurisdiction of an out-of-hospital emergency medical event, under the presence of these shifts, considering as input features a set of clinical variables which evolve over time. Results from our study reveal that, considering the pool of 1 414 575 out-of-hospital medical emergency events from the Valencian region, a dynamic feature domain approach combined with a continual fine-tuning parameter updating strategy stands out as the best option. This approach provides improvements of 4.9% in life-threatening, 18.5% in response delay and 1.7% in jurisdiction, in absolute F1-score, compared to the in-house triage protocol of the Valencian region, and improvements of 4.4% in life-threatening and 11% in response delay, in absolute F1-score, respect to non-continual approaches.

## Data Availability

The data produced in the present work regarding to the performance of the compared strategies is included within the manuscript (figures and tables).

## Acknowledgments

This work has received support from the Ministry of Science, Innovation, and Universities of Spain through the FPU18/06441 program and the KINEMAI project (PID2022-138636OA-I00).

## References

[1] Farand, L., Leprohon, J., Kalina, M., Champagne, F., Contandriopoulos, A.P., Preker, A.: The role of protocols and professional judgement in emergency medical dispatching. European Journal of Emergency Medicine 2(3), 136–148 (1995)

[2] Mackway-Jones, K., Marsden, J., Windle, J.: Emergency triage: Manchester triage group. John Wiley & Sons (2013)

[3] Murray, M., Bullard, M., Grafstein, E.: Revisions to the canadian emergency department triage and acuity scale implementation guidelines. CJEM 6, 421–427 (2004)

[4] Gilboy, N., Tanabe, P., Travers, D.A., Rosenau, A.M., Eitel, D.R. Emergency Severity Index, Version 4: Implementation Handbook. 95. (2012)

[5] Quinonero-Candela, J., Sugiyama, M., Schwaighofer, A., Lawrence, N.D.: Dataset shift in machine learning. MIT Press (2008)

[6] Moreno-Torres, J.G., Raeder, T., Alaiz-Rodríguez, R., Chawla, N.V., Herrera, F.: A unifying view on dataset shift in classification. Pattern Recognition 45(1), 521–530 (2012) 10.1016/j.patcog.2011.06.019

[7] Sáez, C., García-Gómez, J.M.: Kinematics of big biomedical data to characterize temporal variability and seasonality of data repositories: functional data analysis of data temporal evolution over non-parametric statistical manifolds. International journal of medical informatics 119, 109–124 (2018)

[8] Sáez, C., Gutiérrez-Sacristán, A., Kohane, I., García-Gómez, J.M., Avillach, P.: Ehrtemporal-variability: delineating temporal data-set shifts in electronic health records. Gigascience 9(8), 079 (2020)

[9] Guo, L.L., Pfohl, S.R., Fries, J., Posada, J., Fleming, S.L., Aftandilian, C., Shah, N., Sung, L.: Systematic review of approaches to preserve machine learning performance in the presence of temporal dataset shift in clinical medicine. Applied clinical informatics 12(04), 808–815 (2021)

[10] Zhang, A., Xing, L., Zou, J., Wu, J.C.: Shifting machine learning for healthcare from development to deployment and from models to data. Nature Biomedical Engineering 6(12), 1330–1345 (2022)

[11] Ferri, P., Lomonaco, V., Passaro, L.C., Félix-De Castro, A., Sánchez-Cuesta, P., Sáez, C., García-Gómez, J.M.: Deep continual learning for medical call incidents text classification under the presence of dataset shifts. Computers in Biology and Medicine, 108548 (2024)

[12] Ferri, P., Sáez, C., Félix-De Castro, A., Juan-Albarracín, J., Blanes-Selva, V., Sánchez-Cuesta, P., García-Gómez, J.M.: Deep ensemble multitask classification of emergency medical call incidents combining multimodal data improves emergency medical dispatch. Artificial Intelligence in Medicine 117, 102088 (2021)

[13] Guo, L.L., Pfohl, S.R., Fries, J., Johnson, A.E., Posada, J., Aftandilian, C., Shah, N., Sung, L.: Evaluation of domain generalization and adaptation on improving model robustness to temporal dataset shift in clinical medicine. Scientific reports 12(1), 2726 (2022)

[14] Lee, S., Yin, C., Zhang, P.: Stable clinical risk prediction against distribution shift in electronic health records. Patterns 4(9) (2023)

[15] Parisi, G.I., Kemker, R., Part, J.L., Kanan, C., Wermter, S.: Continual lifelong learning with neural networks: A review. Neural Networks 113, 54–71 (2019) 10.1016/j.neunet.2019.01.012

[16] Lomonaco, V., Pellegrini, L., Cossu, A., Carta, A., Graffieti, G., Hayes, T.L., Lange, M., Masana, M., Pomponi, J., Ven, G.M., Mundt, M., She, Q., Cooper, K., Forest, J., Belouadah, E., Calderara, S., Parisi, G.I., Cuzzolin, F., Tolias, A.S., Maltoni, D.: Avalanche: An end-to-end library for continual learning. In: 2021 IEEE/CVF Conference on Computer Vision and Pattern Recognition Workshops (CVPRW, pp. 3595–3605 (2021). 10.1109/CVPRW53098.2021.00399

[17] Nestor, B., McDermott, M.B., Boag, W., Berner, G., Naumann, T., Hughes, M.C., Goldenberg, A., Ghassemi, M.: Feature robustness in non-stationary health records: caveats to deployable model performance in common clinical machine learning tasks. In: Machine Learning for Healthcare Conference, pp. 381–405 (2019). PMLR

[18] Guo, L.L., Steinberg, E., Fleming, S.L., Posada, J., Lemmon, J., Pfohl, S.R., Shah, N., Fries, J., Sung, L.: Ehr foundation models improve robustness in the presence of temporal distribution shift. Scientific Reports 13(1), 3767 (2023)

[19] Lemmon, J., Guo, L.L., Posada, J., Pfohl, S.R., Fries, J., Fleming, S.L., Aftandilian, C., Shah, N., Sung, L.: Evaluation of feature selection methods for preserving machine learning performance in the presence of temporal dataset shift in clinical medicine. Methods of Information in Medicine 62(01/02), 060–070 (2023)

[20] Bengio, Y., Ducharme, R., Vincent, P.: A neural probabilistic language model. Advances in neural information processing systems 13 (2000)

[21] Ho, T.K.: Random decision forests. In: Proceedings of 3rd International Conference on Document Analysis and Recognition, vol. 1, pp. 278–2821 (1995). 10.1109/ICDAR.1995.598994

[22] Friedman, J.H.: Greedy function approximation: A gradient boosting machine. The Annals of Statistics 29(5), 1189–1232 (2001)

[23] Caruana, R.: Multitask learning. Machine learning 28, 41–75 (1997)

[24] Szegedy, C., Vanhoucke, V., Ioffe, S., Shlens, J., Wojna, Z.: Rethinking the inception architecture for computer vision. In: Proceedings of the IEEE Conference on Computer Vision and Pattern Recognition, pp. 2818–2826 (2016)

[25] Rosenblatt, F.: The perceptron: A probabilistic model for information storage and organization in the brain. Psychological Review 65, 386–408 (1958) 10.1037/h0042519

[26] Ba, J.L., Kiros, J.R., Hinton, G.E. Layer Normalization. ArXiv:1607.06450 [Cs, Stat]. (2016). http://arxiv.org/abs/1607.06450

[27] Hendrycks, D., Gimpel, K.: Gaussian error linear units (gelus). arXiv preprint arXiv:1606.08415 (2016)

[28] Hinton, G.E., Srivastava, N., Krizhevsky, A., Sutskever, I., Salakhutdinov, R.R.: Improving neural networks by preventing co-adaptation of feature detectors. arXiv:1207.0580). arXiv. (2012). 10.48550/arXiv.1207.0580. 10.48550/arXiv.1207.0580

[29] Loshchilov, I., Hutter, F. Decoupled Weight Decay Regularization (arXiv:1711.05101). arXiv. (2019). http://arxiv.org/abs/1711.05101

[30] Bertsekas, D.P.: Incremental least squares methods and the extended kalman filter. In: Proceedings of 1994 33rd IEEE Conference on Decision and Control, vol. 2, pp. 1211–1214 (1994). 10.1109/CDC.1994.411166

[31] Janocha, K., Czarnecki, W.M.: On Loss Functions for Deep Neural Networks in Classification. arXiv:1702.05659). arXiv. (2017). http://arxiv.org/abs/1702.05659

[32] Loshchilov, I., Hutter, F.: SGDR: Stochastic Gradient Descent with Warm Restarts. arXiv:1608.03983). arXiv. (2017). 10.48550/arXiv.1608.03983. 10.48550/arXiv.1608.03983

[33] He, K., Zhang, X., Ren, S., Sun, J.: Delving Deep into Rectifiers: Surpassing Human-Level Performance on ImageNet Classification. ArXiv:1502.01852 [Cs]. (2015). http://arxiv.org/abs/1502.01852

[34] Glorot, X., Bengio, Y.: Understanding the difficulty of training deep feedforward neural networks. In: Proceedings of the Thirteenth International Conference on Artificial Intelligence and Statistics, pp. 249–256 (2010). JMLR Workshop and Conference Proceedings

[35] Lan, Z., Chen, M., Goodman, S., Gimpel, K., Sharma, P., Soricut, R.: ALBERT: A Lite BERT for Self-supervised Learning of Language Representations. arXiv:1909.11942). arXiv. (2020). 10.48550/arXiv.1909.11942. 10.48550/arXiv.1909.11942

[36] Face., D.-b.-s.H.: (2023-10-06). https://huggingface.co/dccuchile/albert-base-spanish

[37] Efron, B., Tibshirani, R.J.: An Introduction to the Bootstrap. CRC Press,(1994)

